# Do epigenetic clocks provide explanations for sex differences in lifespan? A cross-sectional twin study

**DOI:** 10.1101/2021.02.23.21252194

**Authors:** Anna Kankaanpää, Asko Tolvanen, Pirkko Saikkonen, Aino Heikkinen, Eija K. Laakkonen, Jaakko Kaprio, Miina Ollikainen, Elina Sillanpää

## Abstract

**Background:** The sex gap in life expectancy has been narrowing in Finland over the past four to five decades; however, on average, women still live longer than men. Epigenetic clocks are markers for biological aging that predict lifespan. In this study, we examined the mediating role of lifestyle factors on the association between sex and biological aging in younger and older adults.

**Methods:** Our sample included same-sex younger and older twins (21-42-y, n = 1110; 50-76-y, n = 763) and younger opposite-sex twins (21-30-y, n = 302). Blood-based DNA methylation (DNAm) was used to compute epigenetic age acceleration by four epigenetic clocks as a measure of biological aging. Path modelling was used to study whether the association between sex and biological aging is mediated through lifestyle-related factors, i.e. education, body mass index, smoking, alcohol use, and physical activity.

**Findings:** In comparison to women, men were biologically older and, in general, they had unhealthier life habits. The effect of sex on biological aging was partly mediated by smoking, but only in older twins. Sex was directly associated with biological aging and the association was stronger in older twins.

**Interpretation:** Previously reported sex differences in lifespan are also evident in biological aging. Declining smoking prevalence among men is a plausible explanation for the narrowing of the difference in life expectancy between the sexes. Data generated by the epigenetic clocks may help in estimating the effects of lifestyle and environmental factors on aging and in predicting aging in future generations.

## INTRODUCTION

Both sexes have experienced tremendous increases in life expectancy over the 20^th^ century. However, through all historical periods, women have had a longer life expectancy than men. The sex gap in life expectancy varies across time and country.^1^ In Finland, the sex gap increased greatly in the first half of the 20^th^ century. That gap was greatest in the mid-1970s (nine years); since then, it has narrowed to 5.4 years.^2^

It has been suggested that sex differences in lifespan are caused by a complex combination of biological (genetic, hormonal) and non-biological (behavioural, economic, social, environmental, and cultural) factors.^3^ Investigating sex differences in cause-specific mortality increases the understanding of the mechanisms underlying the sex differences in overall mortality. In comparison to women, men experience a higher risk of death from almost all causes.^4^ External causes of deaths, such as traffic accidents, trauma, alcohol intoxication, illicit drug overdoses, and suicides, are more common among men. However, at most, these factors typically explain a modest fraction of all premature deaths. The majority of premature deaths are caused by non-communicable diseases (for example, cardiometabolic diseases, lung diseases, cancers, mental disorders, and dementia).^4^ The biological and behavioural factors predisposing an individual to these diseases are predominantly the most important drivers of male-to-female differences in mortality.

Overweight and obesity are dramatically increasing worldwide, predisposing both men and women to several non-communicable diseases.^5^ However, data regarding how obesity trends affect the sex gap in life expectancy are limited. Of the health-hazardous behavioural factors, tobacco smoking has been seen as the predominant driver of both the trend and the extent of sex differences in life expectancy. A recent study suggested that increasing smoking-related mortality among women and decreasing smoking-related mortality among men may account for as much as 40% of the narrowing sex gap in life expectancy over the last two decades.^6^ In general, men consume more alcohol than women. In Finland, the risk for alcohol-related death is three-times higher in men than women.^2^ Globally, men tend to be more physically active than women at all ages,^7^ and leisure-time physical activity is known to be associated with a lower risk of premature death.^8^ Therefore, leisure-time physical activity is expected to diminish the sex gap in life expectancy. Socioeconomic factors might also impact the sex gap, such as differences in education, income, and physical demands of work between the sexes. For example, the sex gap is probably diminishing because many deaths related to trauma and toxication that previously occurred in male-dominated occupations are much rarer nowadays.^5^

In addition to societal factors, differences in innate biology may also have a role in the survival gap between the sexes. Genetic and physiological differences between the sexes include progressive skewing of X chromosome inactivation, telomere attrition, maternally inherited mitochondrial inheritance and hormonal and cellular differences in inflammatory and immunological responses and in substrate metabolism.^9^ The biological longevity advantage of women may also result from oestrogen-associated greater resistance to oxidative damage.^9^ However, in women, sex hormone levels change drastically during menopause, potentially contributing to the reduction of age-related sex differences in health outcomes such as cardiovascular risk factors.

Life expectancy may not always be a reliable proxy for how fast the population is aging, as it is the most distal outcome of ageing processes. To better monitor population health, more sensitive methods are needed to track changes in ageing. Novel biological clocks, i.e. epigenetic clocks, may help track and understand the individual aging process and offer insights into sex differences in biological age and how lifestyle may counteract the aging process.^10–13^ These composite measures have been developed to quantify an individual’s biological age, and they may enable accurate estimation of the pace of aging in all age groups. The first published results on biological age determined by epigenetic clocks have shown that men tend to be biologically older than women.^14–17^

This study aimed to examine sex differences in biological age measured by novel epigenetic clocks in age groups under and over 50 years, with 50 being a proxy for menopausal age.^18^ Moreover, we aimed to assess whether the potential difference in biological aging between the sexes is mediated by different lifestyle factors, and whether age modifies these associations.

## METHODS

### Study population

The Finnish Twin Cohort (FTC) includes three large cohort studies: 1) The older FTC includes twins born before 1958, 2) Finntwin16 includes twins born in 1975-1979, and 3) Finntwin12 includes twins born in 1983-1987.^19–21^ The twins selected from all three cohorts were invited to participate in clinical in-person studies. Data on lifestyle- and health-related traits were collected using questionnaires and interviews. After written informed consent was obtained, blood samples for DNA analyses were collected. Twins (age range from 21 to 76) who had participated in clinical in-person studies with sampling for whole blood DNA and subsequent DNA methylation (DNAm) analyses and who had the relevant phenotype data were included in the current study. Further information on data collection is provided in the appendix (p 1). The analysis sample included monozygotic (MZ) and dizygotic (DZ) same-sex twins (N = 1893, 54% MZ) as well as opposite-sex twin pairs (302 twin individuals).

The FTC data collections were approved by the ethics committees of the University of Helsinki (113/E3/01 and 346/E0/05) and Helsinki University Central Hospital (270/13/03/01/2008 and 154/13/03/00/2011).

## Main variables

### DNA methylation and assessment of biological age

DNAm profiles were obtained using Illumina’s Infinium HumanMethylation450 BeadChip or the Infinium MethylationEPIC BeadChip (Illumina, San Diego, CA, USA). DNAm-based epigenetic age estimates, obtained by Horvath’s^10^ and Hannum’s^11^ clocks and by PhenoAge^12^ and GrimAge^13^ estimators, were calculated using a publicly available online calculator (https://dnamage.genetics.ucla.edu/new). The age acceleration (AA) of each clock was defined as the residual from regressing the estimated biological age on chronological age (AA_Horvath_, AA_Hannum_, AA_Pheno,_ and AA_Grim_, respectively). A more detailed description of the pre-processing and normalising of the DNAm data, as well as additional information on the utilised clocks, is provided in the appendix (pp 1-2).

### Potential mediating variables

We surmised that differences in the covariates between the sex groups are more likely the factors that underlie the sex differences rather than being confounders. The potential mediators included body mass index (BMI), smoking, alcohol use, physical activity, and educational attainment, which is a key component of socio-economic status.

*Educational attainment* was assessed as the number of years of full-time education.

*Body mass index* (BMI), measured as kg/m^2^, can be used as an estimate of healthy diet and sufficient energy intake. A high BMI describes excess fat in the body; thus, it is a consequence of a long-term imbalance between energy intake and expenditure. We measured height in cm using a stadiometer and body mass in kg using a beam scale in kg.

*Smoking* was self-reported and classified as never, former, and current smoker.

*Alcohol use* was measured based on self-reported quantity and frequency of use and the content of the alcohol beverages. These data were transformed into 100% alcohol grams per day.

*Physical activity* was assessed using the Baecke Questionnaire.^22^ The questionnaire has three sections: sports participation, leisure-time physical activity excluding sports, and work- or school-related physical activity. The questionnaire includes four questions on sports activity and leisure-time activity, excluding sports, and eight questions on occupational physical load scored on a five-point scale. A sport index, a non-sport leisure-time (leisure) index, and a work index, respectively, were based on the mean scores of each section as described by Baecke et al.^22^ and Mustelin et al. for the FinnTwin12 study.^23^

### Statistical analysis

To compare differences in the study variables between men and women, we used regression analysis for the continuous variables and the Chi-square test for the categorical variables. In the regression models, the within-pair dependency of twin individuals was taken into account using the cluster option in the analysis.

The shape of the association between age and AA was studied using polynomial models of age as the continuous variable. To study whether sex differences in AA varied by age, the interaction effects of sex and age were also included in the regression models.

Mediation models were used to test whether the association between sex and AA is direct or mediated through lifestyle factors in the same-sex twins and opposite-sex twin pairs. First, the single mediation models were fitted. These models included indirect paths from sex to AA through one lifestyle factor at a time as well as the direct effect of sex on AA. In the same-sex twins, we further studied whether these associations differed according to age group, that is, whether age moderated the associations. The single mediation models included the interaction effect of sex and age group on the mediator variable and directly on AA. Furthermore, the interaction effect between the mediator variable and age group on AA was tested for significance. Second, a multiple mediation model was fitted to assess the mediation effect of the different lifestyle factors simultaneously. The mediators as well as the interactions were included in the final multiple mediation model based on the results of the single mediator models.

The standard errors were corrected for nested sampling using the special option in Mplus (TYPE=COMPLEX). The models for the opposite-sex twin pairs were fitted using multilevel modelling, and the mediation models were specified at the within-twin pairs level. The approach controls for shared childhood environmental factors and partly for genetic factors.

The indirect effects were calculated as the product of the regression coefficients. For the same-sex twins, the age-specific indirect effects were calculated using the parameters of the model. The parameters of the models were estimated using the full information maximum likelihood (FIML) method with robust standard errors. For the models including the ordinal mediator variable smoking status, the estimation was conducted using a robust weighted least squares (WLSMV) estimator. In that case, the mediator is assumed to be the continuous latent variable underlying ordinal smoking status. Descriptive statistics and differences in the study variables were calculated and tested using Stata 16 software (StataCorp, Inc. College Station, TX, USA), and further modelling was conducted with the Mplus statistical package (version 8.2).^24^

## RESULTS

### Sex differences in lifestyle factors and epigenetic aging

The characteristics of the younger and older same-sex twins and the opposite-sex twin pairs included in this study are presented in Table 1. In both age groups of the same-sex twins, there were fewer men than women. Among the older twins, the men were younger and better educated than the women. The men belonging to an opposite-sex twin pair had a lower level of education in comparison to their twin sisters. The men had a higher BMI in young adulthood than the women. Among the same-sex twins, there were more current smokers among the men in comparison to the women; there was no sex difference in smoking among the opposite-sex twin pairs. In all the groups, the men consumed more alcohol than the women. Moreover, the men had a lower level of leisure index in all the groups in comparison to the women, but the men in the younger same-sex twin group had a higher level of sport index than women.

**Table 1.**
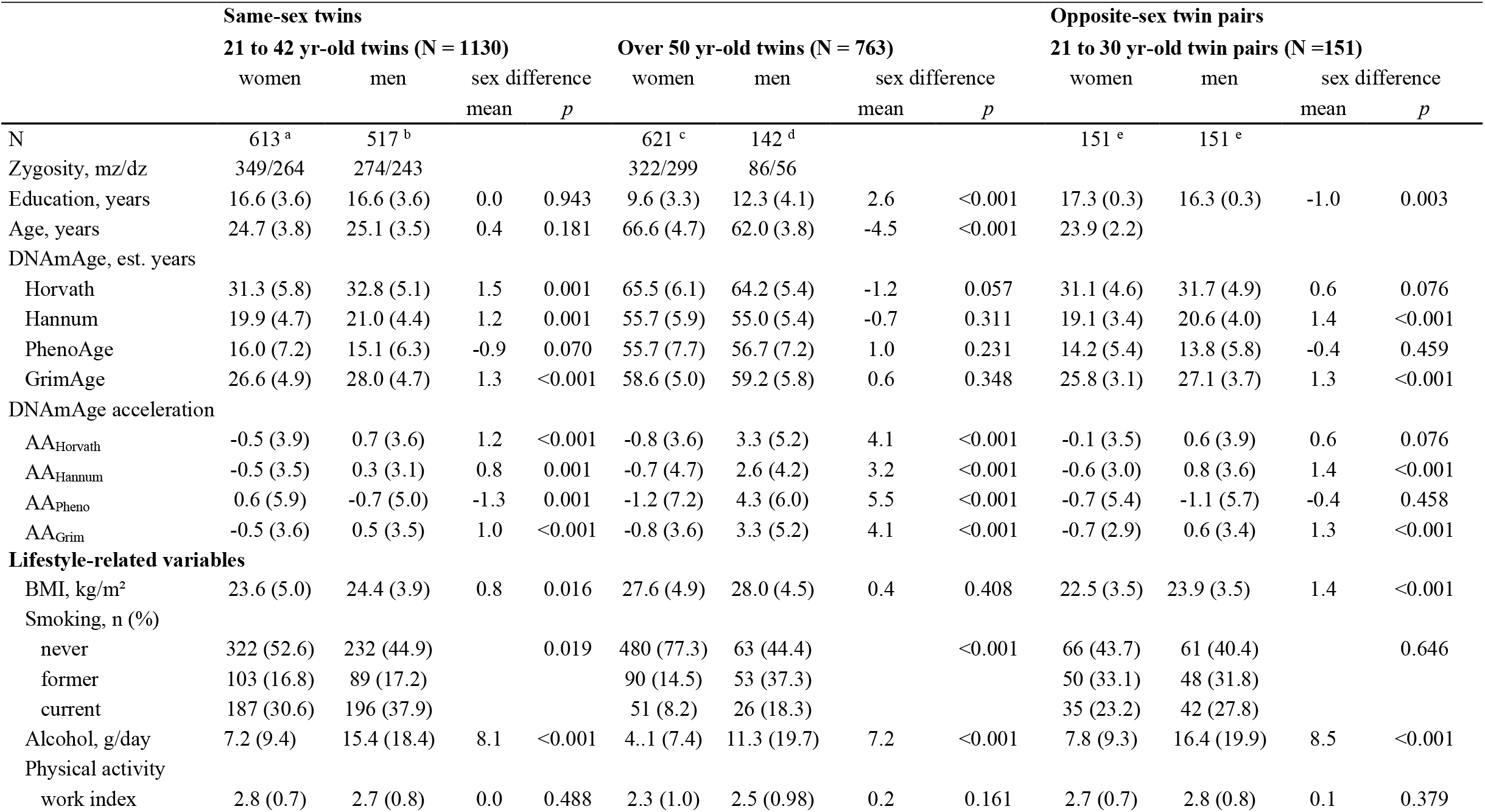
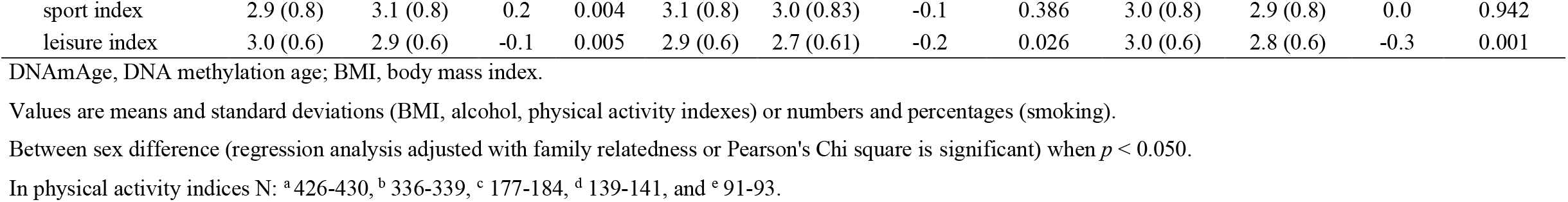
Sex differences in lifestyle-related factors, DNA methylation age and age acceleration (AA) estimates according to age group in the same-sex twins and in opposite-sex twin pairs.

Overall, the men had higher AA than the women, and the sex difference in AA tended to increase with age in the same-sex twins (Table 1 and Figure 1). Interestingly, when DNAm PhenoAge was used to assess AA, the men were epigenetically younger than the women in the younger age, which was in contrast with the AA estimates derived from the other clocks. When controlling for shared childhood environment and partly for genetic factors among the opposite-sex twin pairs, the male twins had higher AA_Hannum_ and AA_Grim_ in comparison to their sisters, but there were no significant differences in AA_Horvath_ and AA_Pheno_ between the sexes (Table 1). A more detailed description of the sex differences in the epigenetic aging is provided in the appendix (pp 2-8).

**Figure 1.**
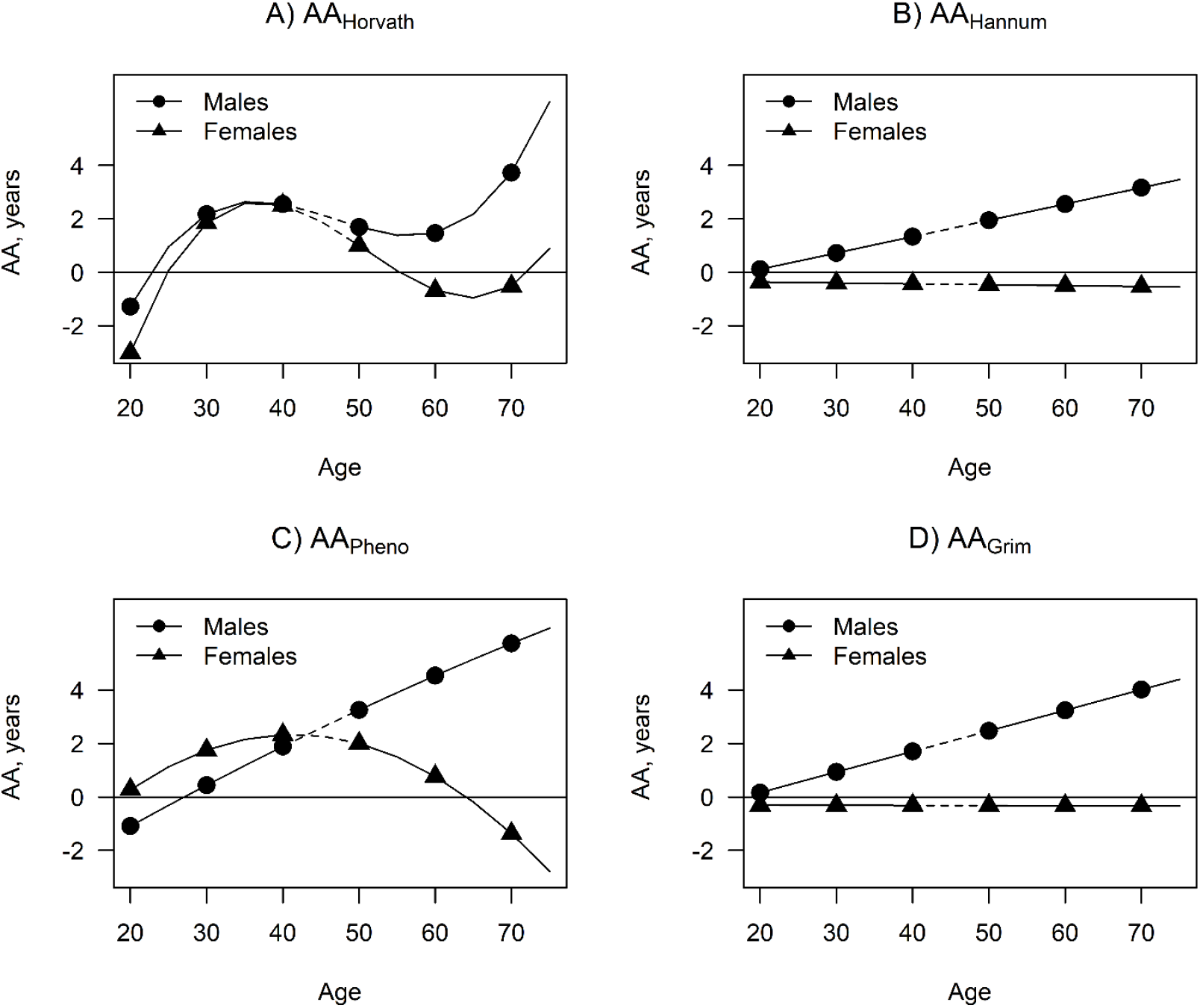
Epigenetic age acceleration (AA) by sex and age in the same-sex twins (n = 1893). Dotted line segments denote the lack of data for 43- to 49-year-olds.

### Mediation models in the twins from the same-sex pairs

Education, BMI, smoking, alcohol use, sport index, leisure index and work index were considered to be the potential mediator variables. Here, smoking was assumed to be a continuous latent variable underlying ordinal smoking status. The estimation results of the single mediator models revealed that the association of sex with education and sport index differed between the age groups (appendix, pp 10-11, mediator regressed on sex × age). There were also some differences in the associations of the potential mediator variables (including education, smoking and alcohol use) with AA between the age groups. The results of the associations between lifestyle-related factors and AA are presented in the appendix (pp 9-11).

The estimated age-specific indirect associations of male sex on AA through the potential mediator variables are shown in Table 2. Male sex was associated with higher AA_Horvath_, AA_Pheno_ and AA_Grim_ through higher BMI, but only in the younger twins. Smoking partly mediated the association of male sex with higher AA_Hannum_, AA_Pheno_ and AA_Grim._ in the older twins. Greater alcohol use partly mediated the association of male sex with higher AA_Pheno_ in the older twins and with AA_Grim_ in both age groups. Moreover, in younger twins, male sex was associated with lower AA_Grim_ through a higher sport index. Because education, leisure index and work index did not mediate the sex difference in AA, these variables were dropped from further modelling.

**Table 2.**
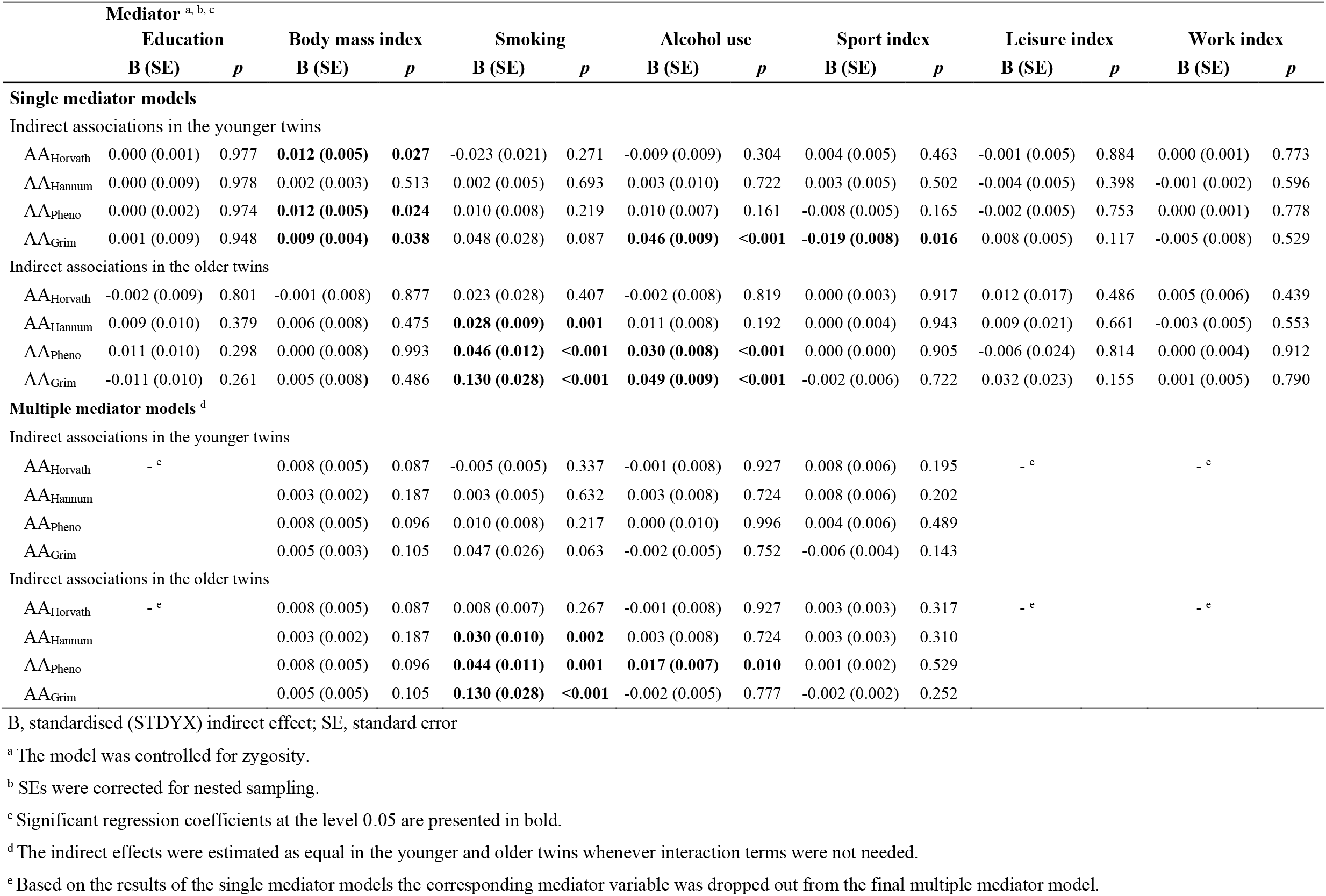
Standardised indirect effects of sex (male) on epigenetic age acceleration (AA) through the potential mediator variables in the same-sex twins.

Based on the results of the single mediator models, the final multiple mediator model included BMI, smoking, alcohol use, and sport index as the mediators (Figure 2). The model also included the interaction effect between sex and age on smoking and sport index, the interaction effect between smoking and age on AA, and the interaction effect between alcohol use and age on AA_Pheno_. When the lifestyle factors were controlled for each other, smoking partly mediated the association between sex and AA (AA_Hannum_, AA_Pheno,_ and AA_Grim_) but only in the older twins (Table 2). Alcohol use partly mediated the sex difference in AA_Pheno_ in the older twins. Male sex was still found to have a direct positive effect on AA_Horvath_, AA_Hannum,_ and AA_Grim_ after all the adjustments, and the association was stronger in the older cohort (Figure 2). Moreover, male sex was found to have a positive direct effect on AA_Pheno_, but only in the older cohort.

**Figure 2.**
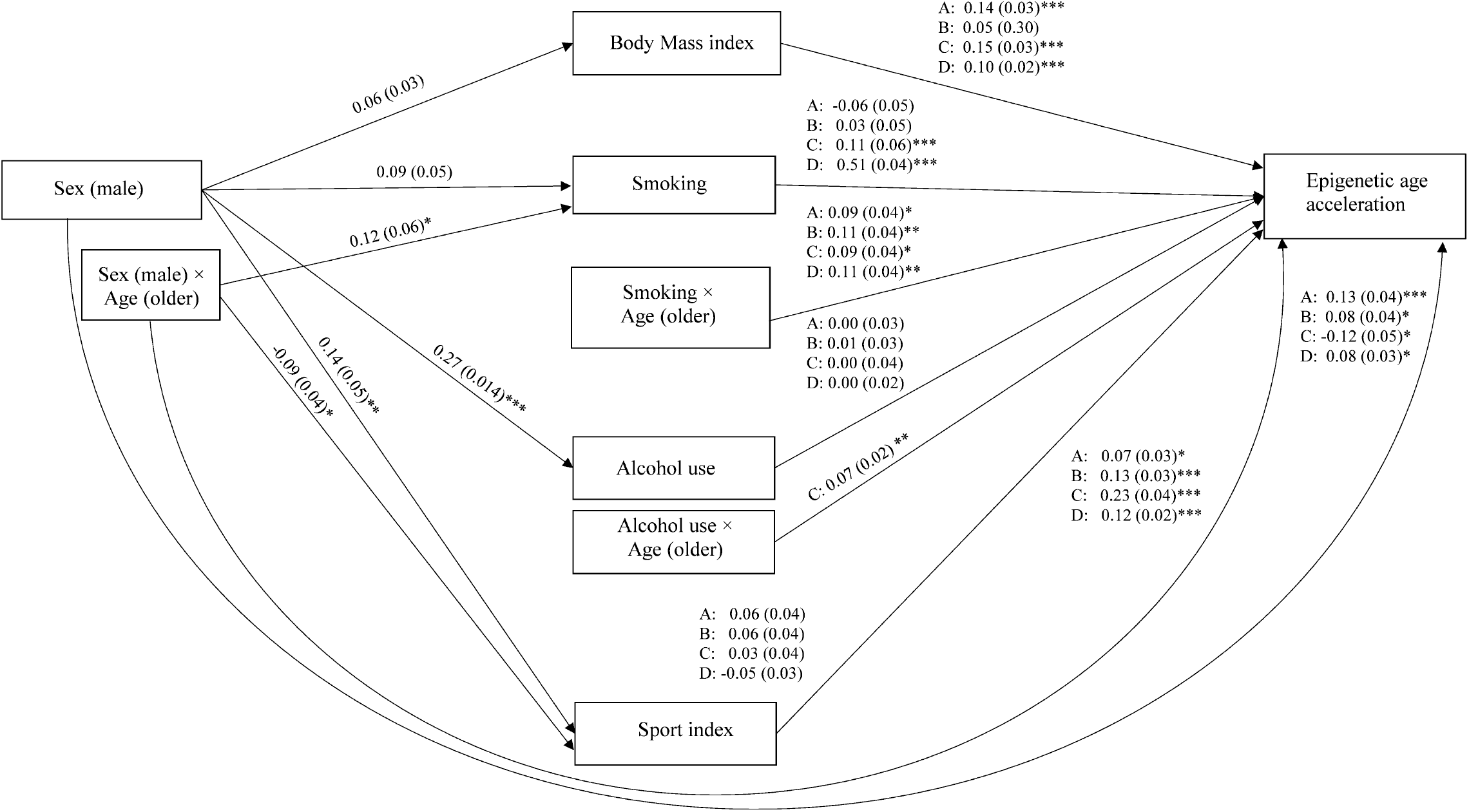
The path diagram of the multiple mediator model in the same-sex twins (n = 1893). Standardised regression coefficients (standard errors) are presented. The modelling was conducted separately for each epigenetic age acceleration (AA) measure: A, AA_Horvath_; B, AA_Hannum_; C, AA_Pheno_; D, AA_Grim_. ***, *p* < 0.001; **, *p* < 0.01; *, *p* < 0.05.

### Mediation models in the opposite-sex twin pairs

Information on the association between the lifestyle-related factors and AA is provided in appendix (pp 12-13). Based on the estimation results of the single mediator models, male sex was associated with accelerated AA_Horvath_ through higher BMI in the opposite-sex twin pairs (Table 3). Otherwise, there were no significant indirect effects. Similar to the models for the same-sex twins, BMI, smoking, alcohol use, and sport index were included in the multiple mediator model as the mediator variables (Figure 3). A significant indirect association of male sex on AA_Horvath_ through higher BMI was also observed after controlling for other lifestyle factors (Table 3). Otherwise, lifestyle factors did not mediate the differences in AA between the men and their female twin sisters. A direct effect of male sex on higher AA_Hannum_ and AA_Grim_ was also observed among the opposite-sex twin pairs (Figure 3).

**Table 3.**
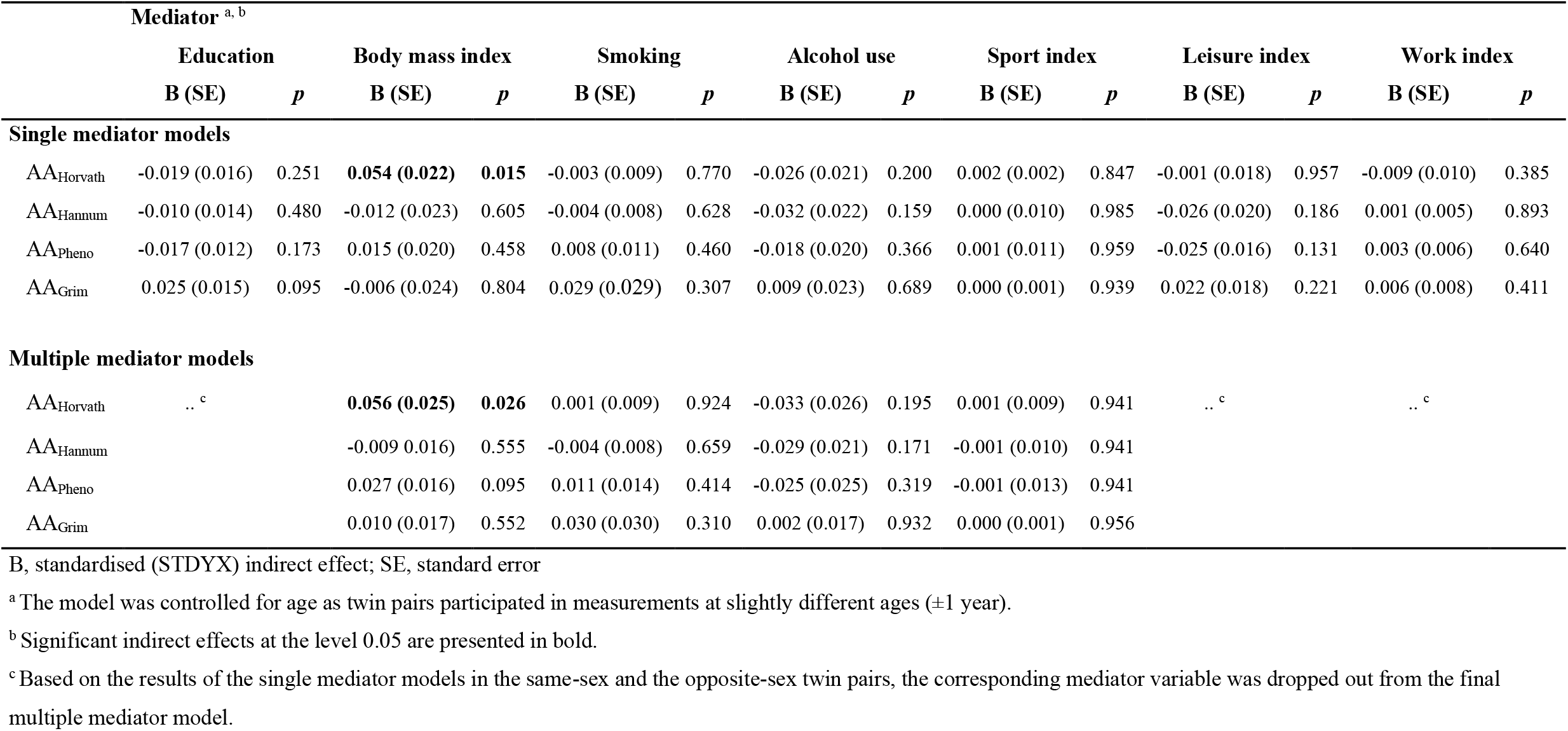
Standardised indirect effects of sex (male) on epigenetic age acceleration (AA) through the potential mediator variables in the opposite-sex twins at within-twin pair level.

**Figure 2.**
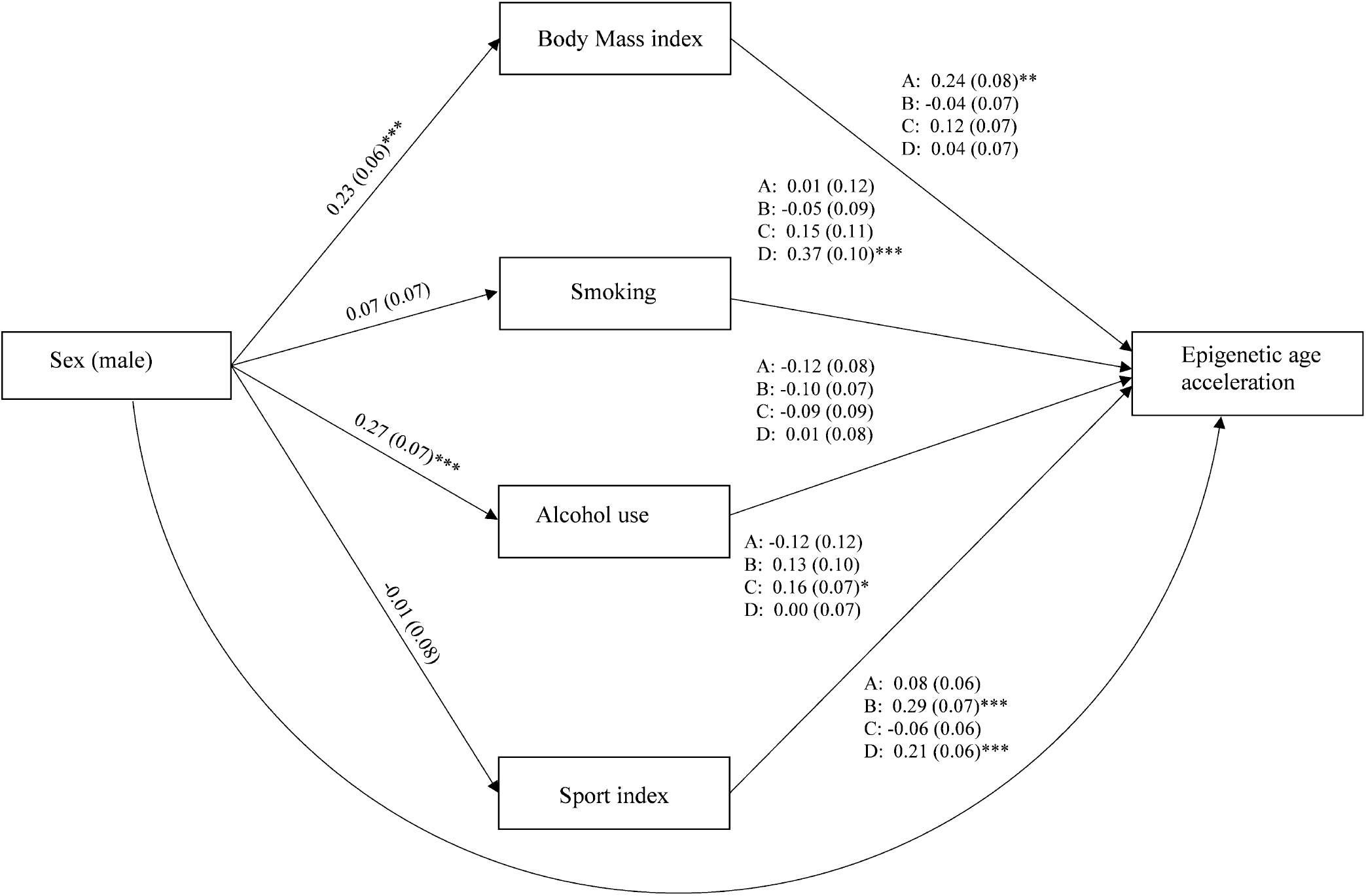
The path diagram of the multiple mediator model in the opposite-sex twin pairs (151 twin pairs). The standardised regression coefficients (standard errors) at the within-twin pair level are presented. The modelling was conducted separately for each epigenetic age acceleration (AA) measure: A, AA_Horvath_; B, AA_Hannum_ C, AA_Pheno_; D, AA_Grim._ ***, *p* < 0.001; **, *p* < 0.01; *, *p* < 0.05.

## DISCUSSION

Our findings suggest that previously reported sex differences in life expectancy can be seen in biological aging when measured with epigenetic clocks (namely Horvath’s clock, Hannum’s clock, DNAm PhenoAge, and GrimAge). Sex difference was already evident in young adulthood and it increased with age; on average, the men were 1.2 to 1.5 years older than women in the younger same-sex twins (21 to 42 years of age) and 3.2 to 4.3 years older in the older same-sex twins (50 to 76 years of age). The only exception was observed in the younger same-sex twins when PhenoAge was used to assess biological aging; the men were epigenetically 1.3 years younger than the women, but the sex difference in biological age reversed in the older adult twins. According to previous studies, sex difference in biological aging measured with epigenetic clocks seems to appear in adolescence,^25^ and men are epigenetically one-to-two years older than women in adulthood.^26^

Opposite-sex twins provide a natural setting for studying sex differences while maximally controlling for genetic factors and shared childhood environmental factors. Epigenetic aging is highly heritable.^27^ Although the share of genes is 50% in male-female dizygotic twins, the mean difference by sex of epigenetic aging within these twin pairs (21 to 30 years of age) was comparable to the sex differences observed in the larger cohort of the same-sex twins.

The observed increase in sex difference with age was mainly due to the fact that epigenetic aging accelerated with age among the men. Changes in the hormonal levels during menopause have a detrimental effect on women’s health,^28^ thus, sex differences in biological aging would be expected to diminish around and after the age of 50. Our analysis studying the shape of the association between chronological age and epigenetic aging did not find any evidence that this is the case in biological aging. This is in line with a recently published study investigating sex differences in the longitudinal trajectories of epigenetic aging from midlife onwards (50 to 90 years of age).^16^ According to the study, men were biologically older than women when Horvath’s and Hannum’s epigenetic clocks and GrimAge were used, and the difference remained constant across the age span.^16^

To the best of our knowledge, this is the first study that has tested the mechanisms underlying sex differences in biological aging measured with epigenetic clocks. We found that several lifestyle-related factors partly mediated the association of sex with biological aging in the same-sex twins, when the mediation of these factors was assessed one at a time. However, after controlling for each of the health-related behaviours in the multiple mediator models, only smoking consistently mediated the sex difference in the older twins. Smoking was associated with accelerated biological aging, and the association was stronger in the older twins; this suggests the cumulative effect of smoking on biological aging. Moreover, the sex difference in smoking behaviour was larger in the older twins; in fact, the difference in the proportion of never-smokers between men and women was wider in the older twins (45% vs 73%) in comparison to the younger twins (45% vs 53%). A previous population-based study investigating long-term trends in smoking in Finland has shown that the prevalence of daily smoking has steadily decreased among men since the late 1970s (37% to 17%); in contrast, the current prevalence among women is about the same as it was four decades ago (∼15%).^29^

Together, these findings support recent studies suggesting that the narrowing of the sex differences in smoking probably partly explains the declining sex gap in lifespan.^6,30^We observed some differences in the associations between lifestyle-related factors and epigenetic aging across the utilised clocks. These inconsistencies are probably due to differences in the procedures used to develop these epigenetic age estimators. The first-generation clocks, namely Horvath’s clock and Hannum’s clock, were trained to predict chronological age. More novel estimators are supposed to also capture CpG sites whose DNAm levels correlate with the deviation of biological age from chronological age.

Of the epigenetic age estimators employed in our study, DNAm GrimAge is the one that has been most recently published, and it outperforms other estimators in terms of predicting mortality.^13,16^ It utilises information on chronological age, sex and seven DNAm-based surrogates for seven plasma proteins and for smoking pack-years. Although sex difference in GrimAge is in-built, reflecting differences in mortality, the observed sex differences were very similar to the corresponding ones measured with Hannum’s clock, which is purely based on CpG sites with their DNAm levels correlating with chronological age. To further understand the sex differences in biological aging, we studied the DNAm-based surrogates included in the GrimAge estimator (see appendix pp 2, 4-8). We observed a significantly higher level of DNAm-based plasminogen activator inhibitor 1 (PAI-1) among men in comparison to women. Moreover, in the men, the level of DNAm PAI-1 drastically increased with age. This DNAm-based surrogate predicts morbidity better than DNAm GrimAge, and it associates with hypertension, type 2 diabetes, and coronary heart disease.^13^ Therefore, higher levels of DNAm PAI-1 in men may play a role in the sex differences in cardiovascular mortality observed in previous studies.^4^

Our study has several strengths. We utilised recently published epigenetic clocks that are shown to predict mortality.^16^ The large sample size of our study enabled us to use complex mediator models. Because data from opposite-sex twin pairs were available, we were also able to control the analyses for shared childhood environmental factors and partly for genetic factors. This study also had some limitations. Most of the studied lifestyle-related factors were self-reported. Furthermore, our data were cross-sectional, and our analysis did not rule out the possibility of reversed causality when studying the associations between lifestyle-related factors and epigenetic aging.

Our results deepen the understanding of the association between sex-dependent lifestyle factors and the aging process. Progress in this area may result in methods that are able to determine individual trajectories in aging that already occur in early adulthood. These methods would enable researchers to investigate the effects of environmental and societal changes and lifestyle interventions on biological aging.

## Data Availability

The dataset used in the current study will be located in the Biobank of the National Institute for Health and Welfare, Finland. All the biobanked data are publicly available for use by qualified researchers following a standardised application procedure

## Contributors

ES conceived the idea for the study. AH and AK pre-processed the DNAm data. AK, ES, and AH accessed and verified the dataset. AK, AT, and ES designed the statistical analysis, and AK and ES performed the analyses. JK and MO have designed and collected the FTC dataset and participated in designing the analysis of this manuscript. AK and ES drafted the first version of the manuscript. All authors contributed to the interpretation of results and drafting or revising the manuscript.

## Declaration of interests

The authors declare that there are no conflicts of interest.

## Acknowledgements

This work was supported by the Academy of Finland (213506, 265240, 263278, 312073 to JK, and 297908 to MO), EC FP5 GenomEUtwin (JK), National Institutes of Health/National Heart, Lung, and Blood Institute (grant HL104125), EC MC ITN Project EPITRAIN (JK and MO), the University of Helsinki Research Funds (MO), Sigrid Juselius Foundation (JK and MO), Yrjö Jahnsson Foundation (6868), and Juho Vainio Foundation (ES).

The authors thank all the FTC study participants and research team members. The Gerontology Research Center is a joint effort between the University of Jyväskylä and the University of Tampere.

## Supplementary Material

Supplementary appendix

## Notes

### Competing Interest Statement

The authors have declared no competing interest.

### Author Declarations

The FTC data collections were approved by the ethics committees of the University of Helsinki (113/E3/01 and 346/E0/05) and Helsinki University Central Hospital (270/13/03/01/2008 and 154/13/03/00/2011). Written informed consent was provided by the participants before the sample collection.

