## Supplementary appendix for "Do epigenetic clocks provide explanations for sex differences in lifespan? A cross-sectional twin study"

**Methods**

The older Finnish Twin Cohort (FTC) was established 45 years ago and data collection has been extensively described recently.^1^ Finntwin16 was initiated in 1991 and to date, it includes five waves of completed data collections.^2^ The main scope of the project is to identify the genetic and environmental determinants of various health-related behaviors and diseases in different stages of life. Finntwin12 is the youngest of the three FTC cohorts.^3^ All eligible twins born in Finland during 1983–1987 along with their biological parents were enrolled to participate to four waves of questionnaires. Selected twins took part in laboratory studies with repeated interviews, neuropsychological tests, and collection of DNA were made as part of wave 4 in early adulthood.^3^ Zygosity of same-sex pairs was confirmed by multiple genetic markers from genome-wide array data.

**Sample collection and DNA methylation analysis**

Genomic DNA was extracted from peripheral blood samples using commercial kits. High molecular weight DNA samples (1 µg) were bisulfite converted using EZ-96 DNA methylation-Gold Kit (Zymo Research, Irvine, CA, USA) according to the manufacturer's protocol. The twins and co-twins were randomly distributed across plates, with both twins from a pair on the same plate. DNA methylation (DNAm) profiles were obtained using Illumina’s Infinium HumanMethylation450 BeadChip or the Infinium MethylationEPIC BeadChip (Illumina, San Diego, CA, USA). The Illumina BeadChips measure single-CpG resolution DNAm levels across the human genome. With these assays, it is possible to interrogate over 450,000 (450k) or 850,000 (EPIC) methylation sites quantitatively across the genome at single-nucleotide resolution. Methylation data were preprocessed using R package *minfi*. Detection *p* values comparing total signal for each probe to the background signal level, were calculated to evaluate quality of the samples.^4^ Samples of poor quality (mean detection *p* > 0.01) were excluded from further analysis. Data were normalized by using the single-sample Noob normalization method, which is suitable for datasets originating from different platforms.^5^ Beta values representing CpG methylation levels were calculated as ratio of methylated intensities (M) to the overall intensities (Beta value=M/(M+U+100), where U is unmethylated probe intensity). β values of the selected probes were used as the input to calculate the predicted age by different epigenetic age estimators using publicly available online calculator (<https://dnamage.genetics.ucla.edu/new>).

**Assessment of biological age**

We utilized four epigenetic clocks to produce biological age estimates. Horvath’s and Hannum’s versions incorporate methylation levels of 353 and 71 age-related CpGs, respectively, and were trained via regressing on chronological age through a penalized regression model.^6,7^ The third epigenetic age estimator, DNAm PhenoAge, was trained on a composite clinical measure of phenotypic age and includes 513 CpG sites.^8^ The newest epigenetic clock, DNAm GrimAge, includes 1030 CpG sites^9^ and was a product of the two-step development method. It first utilized DNAm data to predict a set of biomarkers (plasma proteins and smoking pack-year) and then these developed DNAm-based biomarkers were used to predict mortality.^9^ In both steps, information on participant’s sex and chronological age was used as well.

Age acceleration (AA) of each clock was defined as the residual resulting from regressing predicted biological age on chronological age. By definition AA is not related to chronological age and can be used as a proxy of lower or accelerated epigenetic aging. AA was calculated separately using epigenetic age estimates obtained by Horvath’s clock, Hannum’s clock, DNAm PhenoAge, and DNAm GrimAge estimators (AA_Horvath_, AA_Hannum_, AA_Pheno,_ and AA_Grim_, respectively).

The components of DNAm GrimAge (adjusted for age) were obtained as well, including DNAm-based smoking pack-years (PACYRS) and the surrogates for plasma proteins (DNAm-based plasma proteins): DNAm adrenomedullin (ADM), DNAm beta-2-microglobulin (B2M), DNAm cystatin C, DNAm growth differentiation factor 15 (GDF15), DNAm leptin, DNAm plasminogen activator inhibitor 1 (PAI-1) and DNAm tissue inhibitor metalloproteinases 1 (TIMP-1).

**Results**

**Sex differences in epigenetic aging**

Among the same-sex twins, the shape of the association between age and AA was studied using polynomial (cubic, quadratic and linear) models of age (appendix Table 1). The polynomial with the highest order term and significant regression coefficients was considered optimal. To study whether sex differences in age acceleration (AA) varied by age, the interaction effects of sex and age (Sex **× Age^3^**, Sex **× Age^2^** and Sex **× Age) were also included in the regression models.** For AA_Horvath_ a third order term (Age^3^) in cubic model was significant suggesting that the association between age and AA_Horvath_ followed a cubic curve. There was a difference in the second order terms between the sexes (Sex × Age^2^). The men were biologically older than the women at younger age, but the difference disappeared after 30 years of age and widened again after 50 years of age (Figure 1). For AA_Pheno_ a second order term was significant and therefore, the quadratic model was chosen (appendix Table 1). There was a difference in a linear term between sexes (Sex × Age). Men had lower level of AA_Pheno_ at younger age, but the difference was opposite and increased steeply after 50 years of age (Figure 1). The linear model was sufficient for AA_Hannum_ and AA_Grim_ (appendix Table 1). The men had higher level of AA_Hannum_ and AA_Grim_, and the difference increased with advancing age (Figure 1 and appendix Table 1).

| **Appendix Table 1. The association between age and epigenetic age acceleration (AA) modelled as a third to first order polynomial function of age.** | | | | | | | | | |
| --- | --- | --- | --- | --- | --- | --- | --- | --- | --- |
|  | **Cubic model ^a,b^** | | | **Quadratic model ^a,b^** | | | **Linear model ^a,b^** | | |
|  | **B** | **SE** | ***p*** | **B** | **SE** | ***p*** | **B** | **SE** | ***p*** |
| Sex (male) |  |  |  |  |  |  |  |  |  |
| AA_Horvath_ | 0.01 | 0.08 | 0.921 | ..^c^ |  |  | ..^c^ |  |  |
| AA_Hannum_ | 0.16 | 0.09 | 0.065 | 0.18 | 0.08 | 0.027 | 0.22 | 0.03 | <0.001 |
| AA_Pheno_ | -0.04 | 0.08 | 0.638 | -0.02 | 0.08 | 0.786 | ..^c^ |  |  |
| AA_Grim_ | 0.33 | 0.10 | 0.001 | 0.37 | 0.09 | <0.001 | 0.29 | 0.03 | <0.001 |
| Age |  |  |  |  |  |  |  |  |  |
| AA_Horvath_ | -0.51 | 0.11 | <0.001 | ..^c^ |  |  | ..^c^ |  |  |
| AA_Hannum_ | -0.03 | 0.12 | 0.827 | 0.02 | 0.05 | 0.705 | -0.02 | 0.04 | 0.595 |
| AA_Pheno_ | -0.06 | 0.12 | 0.606 | 0.00 | 0.05 | 0.971 | ..^c^ |  |  |
| AA_Grim_ | -0.11 | 0.11 | 0.314 | 0.01 | 0.05 | 0.858 | -0.03 | 0.03 | 0.264 |
| Age^2^ |  |  |  |  |  |  |  |  |  |
| AA_Horvath_ | -0.54 | 0.08 | <0.001 | ..^c^ |  |  | ..^d^ |  |  |
| AA_Hannum_ | -0.08 | 0.08 | 0.304 | -0.05 | 0.05 | 0.374 |  |  |  |
| AA_Pheno_ | -0.21 | 0.08 | 0.008 | -0.17 | 0.05 | 0.001 |  |  |  |
| AA_Grim_ | -0.13 | 0.08 | 0.110 | -0.05 | 0.05 | 0.274 |  |  |  |
| Age^3^ |  |  |  |  |  |  |  |  |  |
| AA_Horvath_ | 0.98 | 0.15 | <0.001 | ..^d^ |  |  | ..^d^ |  |  |
| AA_Hannum_ | 0.08 | 0.16 | 0.641 |  |  |  |  |  |  |
| AA_Pheno_ | 0.10 | 0.15 | 0.515 |  |  |  |  |  |  |
| AA_Grim_ | 0.19 | 0.15 | 0.207 |  |  |  |  |  |  |
| Sex × Age |  |  |  |  |  |  |  |  |  |
| AA_Horvath_ | 0.08 | 0.11 | 0.465 | ..^c^ |  |  | ..^c^ |  |  |
| AA_Hannum_ | 0.15 | 0.13 | 0.236 | 0.14 | 0.04 | <0.001 | 0.16 | 0.03 | <0.001 |
| AA_Pheno_ | 0.33 | 0.11 | 0.003 | 0.22 | 0.04 | <0.001 | ..^c^ |  |  |
| AA_Grim_ | 0.36 | 0.13 | 0.006 | 0.22 | 0.05 | <0.001 | 0.21 | 0.04 | <0.001 |
| Sex × Age^2^ |  |  |  |  |  |  |  |  |  |
| AA_Horvath_ | 0.19 | 0.08 | 0.019 | ..^c^ |  |  | ..^d^ |  |  |
| AA_Hannum_ | 0.05 | 0.08 | 0.505 | 0.03 | 0.07 | 0.640 |  |  |  |
| AA_Pheno_ | 0.15 | 0.08 | 0.057 | 0.11 | 0.07 | 0.104 |  |  |  |
| AA_Grim_ | -0.02 | 0.09 | 0.846 | -0.09 | 0.08 | 0.266 |  |  |  |
| Sex × Age^3^ |  |  |  |  |  |  |  |  |  |
| AA_Horvath_ | -0.03 | 0.10 | 0.802 | ..^d^ |  |  | ..^d^ |  |  |
| AA_Hannum_ | -0.01 | 0.11 | 0.952 |  |  |  |  |  |  |
| AA_Pheno_ | -0.12 | 0.11 | 0.279 |  |  |  |  |  |  |
| AA_Grim_ | -0.14 | 0.12 | 0.217 |  |  |  |  |  |  |
| B, standardised regression coefficient; SE, standard error. | | | | |  |  |  |  |  |
| ^a^ The model was controlled for zygosity. | | | | | | | | | |
| ^b^ SEs were corrected for nested sampling. | | | | | | | | | |
| ^c^ The model was not fitted, because a higher order polynomial model was needed for the AA measure. | | | | | | | | | |
| ^d^ The model did not include the corresponding polynomial term. | | | | | | | | | |

**Sex differences in DNAm-based plasma proteins and smoking pack-years**

Information on sex is utilized in the estimation of epigenetic age by GrimAge estimator.^9^ Therefore, the observed sex difference in AA_Grim_ may reflect the estimated sex difference in mortality and not only the differences in DNAm. Also, in DNAm-based surrogates included in the GrimAge estimator the sex difference is in-built, indicating differences between men and women in actual levels of plasma proteins and smoking pack-years. To further understand the sex differences in biological aging, we also studied the differences in in age-adjusted DNAm-based plasma proteins and smoking pack-years in the same-sex twins and in the opposite-sex twin pairs. The variables were standardized before the analysis.

For DNAm-based ADM and B2M, a linear model was sufficient (appendix Table 2). Men had a lower level of DNAm ADM and B2M in young adulthood, but the difference narrowed or disappeared with age (appendix Figure 1, A‒B). For DNAm GDF15, a quadratic model was required, and there was a sex difference in the second order term (appendix Table 2). Among women the association followed U-shaped pattern, whereas among men the level of DNAm GDF15 decreased linearly with age (appendix Figure 1, C). The men had slightly lower level of DNAm GDF15 in younger age. The difference disappeared after 30 years of age but widened again after 50 years of age. Association between age and DNAm cystatin C followed a quadratic curve, and there was a sex difference in the linear term (appendix Table 2). The sex difference in this surrogate increased rather steeply from midlife onwards and the men had a higher level especially in older age (appendix Figure 1, D). For DNAm leptin the difference in a third order term between the sexes was significant in a cubic model (appendix Table 2). Overall, the men had a lower level of DNAm leptin (appendix Figure 1, E). In younger age, the sex difference was constant across ages but after 50 years of age the difference slightly narrowed. For DNAm PAI-1 a quadratic model was required (appendix Table 2). Overall, the men had higher level of DNAm PAI-1, and the sex difference increased rather steeply with age (appendix Figure 1, F). For DNAm TIMP-1 and DNAm packyrs a linear model was sufficient (appendix Table 2). The sex difference in DNAm TIMP-1 increased with age and the men had a higher level of DNAm TIMP-1 especially in older age (appendix Figure 1, G). The men had also a higher level of DNAm-based smoking pack-years, and the difference did not depend on age (appendix Figure 1, H).

| **Appendix Table 2. The association between age and the standardised age-adjusted DNA methylation (DNAm)-based plasma proteins and smoking pack-years modelled as a third to first order polynomial function of age (n = 1893).** | | | | | | | | | |
| --- | --- | --- | --- | --- | --- | --- | --- | --- | --- |
|  | **Cubic model ^a,b^** | | | **Quadratic model ^a,b^** | | | **Linear model ^a,b^** | | |
|  | **B** | **SE** | ***p*** | **B** | **SE** | ***p*** | **B** | **SE** | ***p*** |
| Sex (male) |  |  |  |  |  |  |  |  |  |
| DNAm ADM | -0.67 | 0.08 | <0.001 | -0.62 | 0.07 | <0.001 | -0.68 | 0.02 | <0.001 |
| DNAm B2M | 0.01 | 0.10 | 0.944 | 0.05 | 0.09 | 0.557 | -0.09 | 0.04 | 0.014 |
| DNAm GDF15 | -0.06 | 0.08 | 0.421 | 0.00 | 0.07 | 0.966 | .. ^c^ |  |  |
| DNAm cystatin C | 0.04 | 0.10 | 0.736 | 0.08 | 0.09 | 0.372 | .. ^c^ |  |  |
| DNAm leptin | -0.93 | 0.04 | <0.001 | .. ^c^ |  |  | .. ^c^ |  |  |
| DNAm PAI-1 | 0.26 | 0.08 | 0.001 | 0.22 | 0.07 | 0.003 | .. ^c^ |  |  |
| DNAn TIMP-1 | 0.16 | 0.09 | 0.072 | 0.23 | 0.08 | 0.004 | 0.16 | 0.03 | <0.001 |
| DNAm packyrs | 0.26 | 0.11 | 0.020 | 0.24 | 0.10 | 0.020 | 0.11 | 0.03 | 0.002 |
| Age |  |  |  |  |  |  |  |  |  |
| DNAm ADM | -0.40 | 0.08 | <0.001 | -0.26 | 0.04 | <0.001 | -0.27 | 0.02 | <0.001 |
| DNAm B2M | -0.26 | 0.11 | 0.022 | -0.13 | 0.05 | 0.016 | -0.09 | 0.03 | 0.010 |
| DNAm GDF15 | -0.33 | 0.12 | 0.005 | -0.20 | 0.05 | <0.001 | .. ^c^ |  |  |
| DNAm cystatin C | -0.34 | 0.11 | 0.002 | -0.19 | 0.06 | 0.001 | .. ^c^ |  |  |
| DNAm leptin | -0.25 | 0.06 | <0.001 | .. ^c^ |  |  | .. ^c^ |  |  |
| DNAm PAI-1 | 0.25 | 0.13 | 0.046 | 0.14 | 0.05 | 0.007 | .. ^c^ |  |  |
| DNAn TIMP-1 | -0.28 | 0.10 | 0.005 | -0.12 | 0.06 | 0.037 | -0.07 | 0.04 | 0.063 |
| DNAm packyrs | 0.04 | 0.10 | 0.734 | -0.04 | 0.05 | 0.459 | 0.03 | 0.03 | 0.303 |
| Age^2^ |  |  |  |  |  |  |  |  |  |
| DNAm ADM | -0.12 | 0.07 | 0.074 | -0.02 | 0.04 | 0.579 | ..^d^ |  |  |
| DNAm B2M | -0.04 | 0.09 | 0.651 | 0.17 | 0.05 | 0.294 |  |  |  |
| DNAm GDF15 | 0.16 | 0.07 | 0.022 | 0.25 | 0.05 | <0.001 |  |  |  |
| DNAm cystatin C | 0.04 | 0.09 | 0.687 | 0.14 | 0.05 | 0.006 |  |  |  |
| DNAm leptin | -0.16 | 0.04 | <0.001 | .. ^c^ |  |  |  |  |  |
| DNAm PAI-1 | -0.11 | 0.08 | 0.185 | -0.18 | 0.06 | 0.001 |  |  |  |
| DNAn TIMP-1 | -0.04 | 0.09 | 0.666 | 0.07 | 0.05 | 0.148 |  |  |  |
| DNAm packyrs | 0.13 | 0.09 | 0.148 | 0.08 | 0.05 | 0.102 |  |  |  |
| Age^3^ |  |  |  |  |  |  |  |  |  |
| DNAm ADM | 0.23 | 0.12 | 0.053 | ..^d^ |  |  | ..^d^ |  |  |
| DNAm B2M | 0.20 | 0.16 | 0.200 |  |  |  |  |  |  |
| DNAm GDF15 | 0.20 | 0.14 | 0.146 |  |  |  |  |  |  |
| DNAm cystatin C | 0.23 | 0.16 | 0.134 |  |  |  |  |  |  |
| DNAm leptin | 0.06 | 0.08 | 0.488 |  |  |  |  |  |  |
| DNAm PAI-1 | -0.17 | 0.16 | 0.288 |  |  |  |  |  |  |
| DNAn TIMP-1 | 0.25 | 0.14 | 0.084 |  |  |  |  |  |  |
| DNAm packyrs | -0.11 | 0.15 | 0.440 |  |  |  |  |  |  |
| Sex × Age |  |  |  |  |  |  |  |  |  |
| DNAm ADM | -0.20 | 0.10 | 0.051 | 0.17 | 0.03 | <0.001 | 0.16 | 0.03 | <0.001 |
| DNAm B2M | 0.32 | 0.16 | 0.046 | 0.17 | 0.05 | <0.001 | 0.13 | 0.04 | 0.001 |
| DNAm GDF15 | -0.17 | 0.09 | 0.074 | 0.00 | 0.03 | 0.829 | .. ^c^ |  |  |
| DNAm cystatin C | 0.49 | 0.15 | 0.001 | 0.25 | 0.05 | <0.001 | .. ^c^ |  |  |
| DNAm leptin | -0.03 | 0.06 | 0.664 | .. ^c^ |  |  | .. ^c^ |  |  |
| DNAm PAI-1 | 0.10 | 0.11 | 0.368 | 0.15 | 0.04 | <0.001 | .. ^c^ |  |  |
| DNAn TIMP-1 | 0.27 | 0.11 | 0.020 | 0.26 | 0.05 | <0.001 | 0.23 | 0.04 | <0.001 |
| DNAm packyrs | -0.16 | 0.14 | 0.556 | -0.01 | 0.05 | 0.913 | -0.05 | 0.04 | 0.201 |
| Sex × Age^2^ |  |  |  |  |  |  |  |  |  |
| DNAm ADM | 0.00 | 0.07 | 0.968 | -0.08 | 0.06 | 0.226 | ..^d^ |  |  |
| DNAm B2M | -0.06 | 0.21 | 0.491 | -0.34 | 0.07 | 0.052 |  |  |  |
| DNAm GDF15 | -0.14 | 0.07 | 0.055 | -0.17 | 0.06 | 0.008 |  |  |  |
| DNAm cystatin C | 0.11 | 0.09 | 0.214 | 0.01 | 0.07 | 0.876 |  |  |  |
| DNAm leptin | 0.01 | 0.04 | 0.912 | .. ^c^ |  |  |  |  |  |
| DNAm PAI-1 | 0.05 | 0.08 | 0.489 | 0.11 | 0.07 | 0.099 |  |  |  |
| DNAn TIMP-1 | 0.01 | 0.09 | 0.913 | -0.06 | 0.07 | 0.398 |  |  |  |
| DNAm packyrs | -0.16 | 0.11 | 0.135 | -0.12 | 0.09 | 0.188 |  |  |  |
| Sex × Age^3^ |  |  |  |  |  |  |  |  |  |
| DNAm ADM | -0.04 | 0.09 | 0.682 | ..^d^ |  |  | ..^d^ |  |  |
| DNAm B2M | -0.15 | 0.14 | 0.264 |  |  |  |  |  |  |
| DNAm GDF15 | 0.16 | 0.09 | 0.078 |  |  |  |  |  |  |
| DNAm cystatin C | -0.25 | 0.13 | 0.059 |  |  |  |  |  |  |
| DNAm leptin | 0.12 | 0.06 | 0.034 |  |  |  |  |  |  |
| DNAm PAI-1 | 0.06 | 0.11 | 0.561 |  |  |  |  |  |  |
| DNAn TIMP-1 | -0.01 | 0.11 | 0.915 |  |  |  |  |  |  |
| DNAm packyrs | 0.08 | 0.13 | 0.532 |  |  |  |  |  |  |
| ADM, adrenomedullin; B2M, beta-2 microglobulin; GDF15, growth differentiation factor 15; PAI-1, plasminogen activation inhibitor 1; TIMP-1, tissue inhibitor metalloproteinase 1; packyrs, pack-years. | | | | | | | | | |
| B standardised (STDYX) regression coefficient; SE, standard error. | | | | | | | | | |
| ^a^ The model was controlled for zygosity. | | | | | | | | | |
| ^b^ SEs were corrected for nested sampling. | | | | | | | | | |
| ^c^ The model was not fitted, because the higher order polynomial model was needed for the measure. | | | | | | | | | |
| ^d^ The model did not include the corresponding polynomial term. | | | | | | | | | |

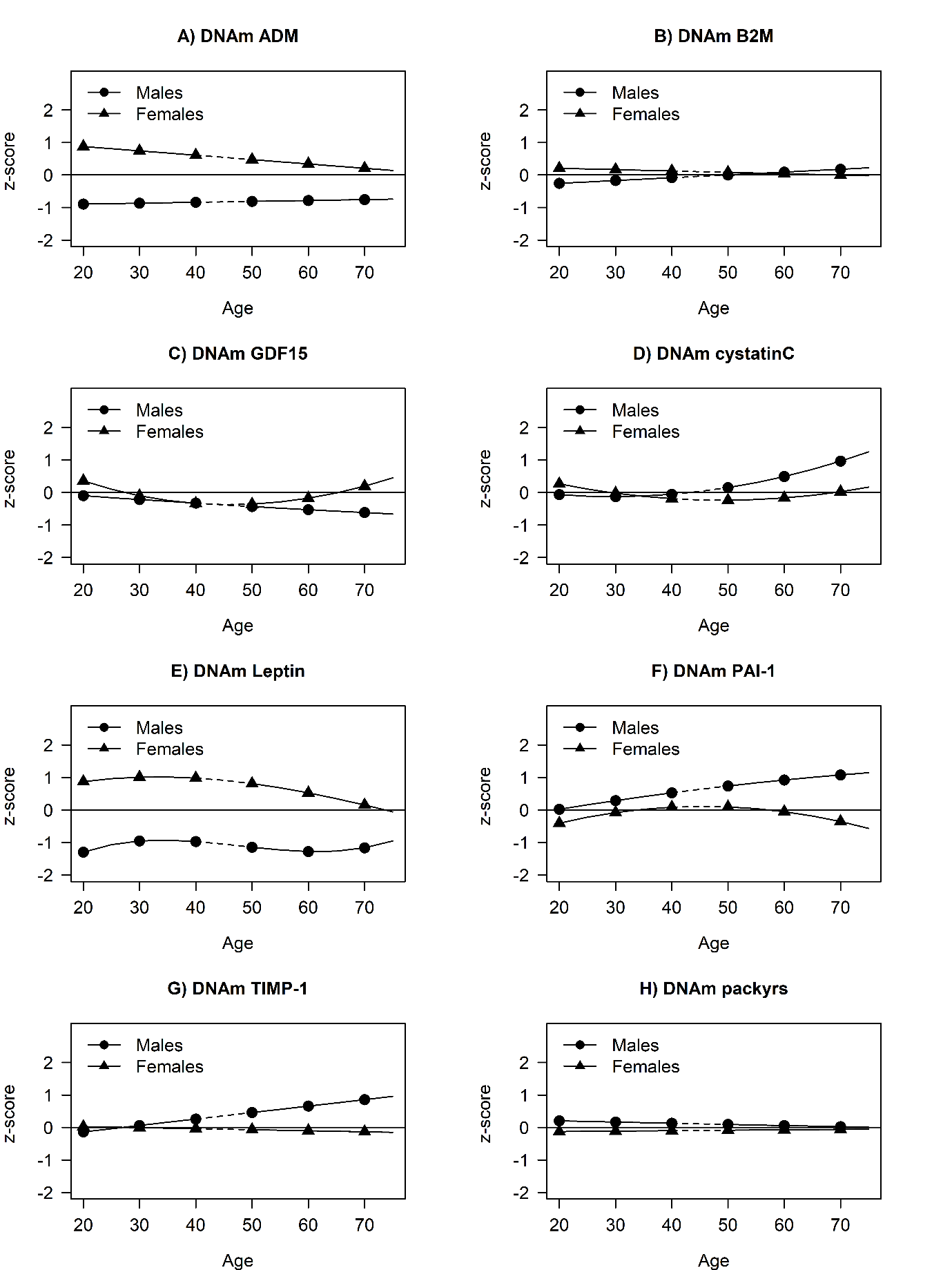

**Appendix Figure 1. DNA methylation (DNAm)-based plasma proteins and smoking pack-years by sex and age in the same-sex twins (n = 1893).**

Dotted line segments denote the lack of data for 43- to 49-year-olds. ADM, adrenomedullin; B2M, beta-2 microglobulin; GDF15, growth differentiation factor 15; PAI-1, plasminogen activation inhibitor 1; TIMP-1, tissue inhibitor metalloproteinase 1; packyrs, pack-years.

In the opposite-sex twin pairs, the men had significantly lower levels of DNAm ADM, B2M, and leptin, whereas higher levels of DNAm PAI-1 and DNAm-based smoking pack-years (appendix Figure 2).
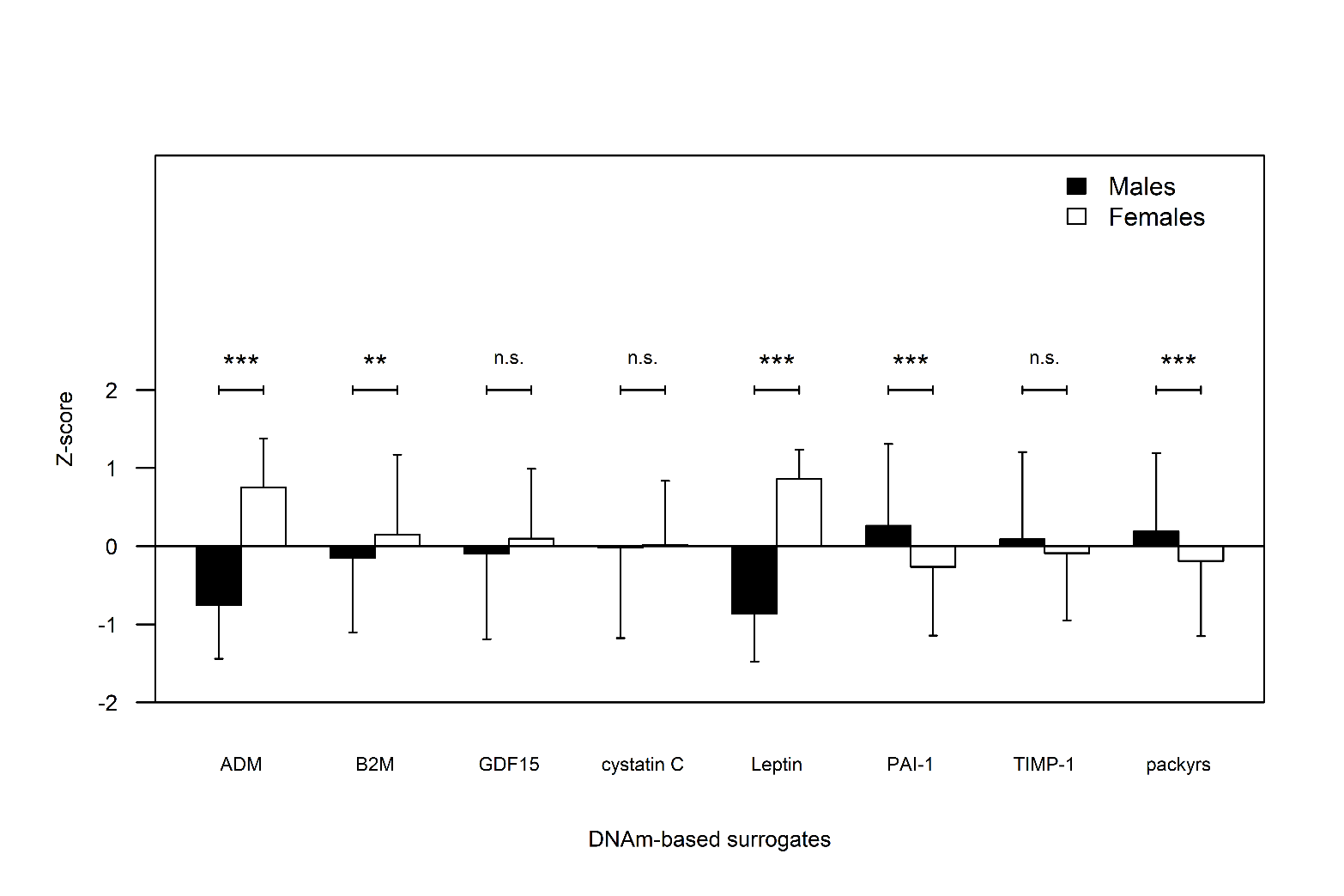

**Appendix Figure 2. The mean levels (and standard deviations) of standardised age-adjusted DNA methylation (DNAm)-based plasma proteins and smoking pack-years by sex in the opposite-sex twin pairs (151 pairs).**

ADM, adrenomedullin; B2M, beta-2 microglobulin; GDF15, growth differentiation factor 15; PAI-1, plasminogen activation inhibitor 1; TIMP-1, tissue inhibitor metalloproteinase 1; packyrs, pack-years;

***, *p* < 0.001; **, *p* < 0.01; *, *p* < 0.05, n.s., not significant; *p-*value for within-twin pair sex difference.

**Mediation models in twins from same-sex pairs**

The associations between lifestyle-related factors and AA estimated using the single mediator models are presented in appendix Table 3. Higher education was associated with slower AA_Pheno_ and AA_Grim_ but only in the younger twins (B=-0.09, education × age: B=0.16 and B=-0.34, education × age: B=0.26, respectively). Larger BMI was associated with accelerated AA_Horvath_ and AA_Pheno_ only in the younger twins, as well (B=0.15., BMI × age: B=-0.17 and B=0.16, BMI × age: B=-0.16, respectively), but the differences in the associations were not significant. Larger BMI was associated with accelerated AA_Grim_ in both age groups (B=0.12). Higher smoking was associated with accelerated AA_Hannum_ only in the older twins (B=0.02, smoking × age B=0.11), whereas smoking was associated with accelerated AA_Pheno_ and AA_Grim_ also in the younger twins and the association was stronger in the older twins (B=0.11, smoking × age: B=0.10, and B=0.53, smoking × age: B=0.10, respectively). Greater alcohol use was associated with accelerated AA_Pheno_, but this was seen only in the older twins (B=0.04; alcohol × age: B=0.08). Furthermore, greater alcohol use was associated with accelerated AA_Grim_ in both age groups (B=0.16). Sport index was associated with slower AA_Grim_ in both age groups (B=-0.17) and work index with higher AA_Grim_ only in the younger twins (B=0.23 and work index × age: B=-0.21, respectively). Leisure index was not associated with AA. In all the models, sex was directly associated with epigenetic aging. Male sex was associated with higher AA_Horvath_, AA_Hannum_ and AA_Grim_. and the association was stronger in the older twins (B=0.08 to 0.15, sex × age: B=0.07 to 0.23). Instead, male sex was associated with slower AA_Pheno_ in the younger twins, but the association turned positive in the older twins (B=-0.11 to -0.09, sex × age: B=0.25 to 0.29).

When the lifestyle factors (BMI, smoking, alcohol use, and sport index) were controlled for each other in the multiple mediator models, higher BMI was still associated with higher AA_Horvath_, AA_Pheno_, and AA_Grim._ Smoking was associated with accelerated AA_Hannum_ only in the older twins (B=0.03 and smoking × age: B=0.11), whereas smoking was associated with accelerated AA_Pheno_ and AA_Grim_ also in the younger twins and the association was stronger in the older twins (B=0.11, smoking × age: B=0.09, and B=0.51, smoking × age: B=0.11, respectively). Greater alcohol use was associated with accelerated AA_Pheno_ only in the older twins (B=0.00; alcohol × age: B=0.07). The direct effect of male sex was still observed on AA_Horvath_, AA_Hannum_ and AA_Grim_, and the association was stronger in the older twins (B=0.08 to 0.13, sex × age: B=0.07 to 0.13). Instead, male sex was associated with slower AA_Pheno_ in the younger twins, but the association turned positive in the older twins (B=-0.12, sex × age: B=0.23).

| **Appendix Table 3. The estimation results of the single mediator models among the same-sex twins (n = 1893).** | | | | | | | | | | | | | | |
| --- | --- | --- | --- | --- | --- | --- | --- | --- | --- | --- | --- | --- | --- | --- |
|  | **Mediator ^a,b,c^** | | | | | | | | | | | | | |
|  | **Education** | | **Body mass index** | | **Smoking** | | **Alcohol use** | | **Sport index** | | **Leisure index** | | **Work index** | |
|  | **B (SE)** | ***p*** | **B (SE)** | ***p*** | **B (SE)** | ***p*** | **B (SE)** | ***p*** | **B (SE)** | ***p*** | **B (SE)** | ***p*** | **B (SE)** | ***p*** |
| **Epigenetic age acceleration regressed on** | | | | | |  |  |  |  |  |  |  |  |  |
| Mediator |  |  |  |  |  |  |  |  |  |  |  |  |  |  |
| AA_Horvath_ | 0.04 (0.04) | 0.363 | **0.15 (0.03)** | **<0.001** | -0.07 (0.05) | 0.183 | -0.03 (0.03) | 0.298 | 0.03 (0.04) | 0.446 | 0.01 (0.04) | 0.884 | 0.02 (0.05) | 0.756 |
| AA_Hannum_ | -0.01 (0.04) | 0.829 | 0.02 (0.03) | 0.503 | 0.02 (0.05) | 0.663 | 0.01 (0.03) | 0.722 | 0.03 (0.04) | 0.492 | 0.04 (0.04) | 0.368 | 0.04 (0.05) | 0.388 |
| AA_Pheno_ | **-0.09 (0.04)** | **0.028** | **0.16 (0.03)** | **<0.001** | **0.11 (0.05)** | **0.020** | 0.04 (0.03) | 0.165 | -0.07 (0.04) | 0.134 | 0.01 (0.05) | 0.752 | 0.02 (0.05) | 0.761 |
| AA_Grim_ | **-0.34 (0.04)** | **<0.001** | **0.12 (0.04)** | **0.002** | **0.53 (0.04)** | **<0.001** | **0.16 (0.03)** | **<0.001** | **-0.17 (0.03)** | **<0.001** | -0.07 (0.04) | 0.080 | **0.23 (0.04)** | **<0.001** |
| Sex (male) |  |  |  |  |  |  |  |  |  |  |  |  |  |  |
| AA_Horvath_ | **0.14 (0.03)** | **<0.001** | **0.12 (0.03)** | **<0.001** | **0.14 (0.04)** | **<0.001** | **0.15 (0.03)** | **<0.001** | **0.13 (0.03)** | **<0.001** | **0.14 (0.03)** | **<0.001** | **0.14 (0.03)** | **<0.001** |
| AA_Hannum_ | **0.10 (0.03)** | **<0.001** | **0.10 (0.03)** | **0.001** | **0.10 (0.04)** | **0.006** | **0.10 (0.03)** | **0.001** | **0.10 (0.03)** | **0.001** | **0.10 (0.03)** | **<0.001** | **0.10 (0.03)** | **<0.001** |
| AA_Pheno_ | **-0.10 (0.03)** | **0.001** | **-0.11 (0.03)** | **<0.001** | **-0.11 (0.05)** | **0.032** | **-0.11 (0.03)** | **<0.001** | **-0.09 (0.03)** | **0.003** | **-0.10 (0.03)** | **0.001** | **-0.10 (0.03)** | **0.001** |
| AA_Grim_ | **0.12 (0.03)** | **<0.001** | **0.12 (0.03)** | **<0.001** | **0.08 (0.03)** | **0.010** | **0.08 (0.03)** | **0.018** | **0.14 (0.03)** | **<0.001** | **0.12 (0.03)** | **<0.001** | **0.13 (0.03)** | **<0.001** |
| Age (older) |  |  |  |  |  |  |  |  |  |  |  |  |  |  |
| AA_Horvath_ | 0.06 (0.10) | 0.584 | 0.09 (0.15) | 0.549 | -0.07 (0.05) | 0.126 | -0.03 (0.04) | 0.398 | 0.02 (0.14) | 0.891 | 0.08 (0.16) | 0.608 | -0.10 (0.11) | 0.395 |
| AA_Hannum_ | -0.07 (0.10) | 0.448 | -0.10 (0.16) | 0.507 | -0.04 (0.05) | 0.428 | -0.01 (0.04) | 0.715 | 0.03 (0.15) | 0.867 | 0.11 (0.20) | 0.574 | 0.09 (0.12) | 0.426 |
| AA_Pheno_ | **-0.32 (0.10)** | **0.001** | -0.02 (0.16) | 0.882 | -0.12 (0.07) | 0.087 | **-0.14 (0.04)** | **<0.001** | -0.20 (0.18) | 0.263 | -0.15 (0.23) | 0.509 | -0.09 (0.12) | 0.447 |
| AA_Grim_ | **-0.50 (0.10)** | **<0.001** | -0.07 (0.15) | 0.648 | **0.11 (0.04)** | **0.005** | -0.02 (0.03) | 0.449 | -0.02 (0.16) | 0.920 | 0.15 (0.19) | 0.448 | **0.22 (0.11)** | **0.044** |
| Sex (male)×Age (older) | |  |  |  |  |  |  |  |  |  |  |  |  |  |
| AA_Horvath_ | **0.08 (0.04)** | **0.016** | **0.09 (0.03)** | **0.011** | **0.07 (0.04)** | **0.050** | **0.08 (0.04)** | **0.028** | **0.09 (0.03)** | **0.014** | **0.08 (0.04)** | **0.022** | **0.08 (0.04)** | **0.025** |
| AA_Hannum_ | **0.15 (0.04)** | **<0.001** | **0.16 (0.04)** | **<0.001** | **0.13 (0.03)** | **<0.001** | **0.15 (0.04)** | **<0.001** | **0.16 (0.04)** | **<0.001** | **0.15 (0.04)** | **<0.001** | **0.16 (0.04)** | **<0.001** |
| AA_Pheno_ | **0.27 (0.04)** | **<0.001** | **0.29 (0.04)** | **<0.001** | **0.25 (0.05)** | **<0.001** | **0.26 (0.04)** | **<0.001** | **0.28 (0.04)** | **<0.001** | **0.28 (0.04)** | **<0.001** | **0.28 (0.04)** | **<0.001** |
| AA_Grim_ | **0.23 (0.04)** | **<0.001** | **0.21 (0.04)** | **<0.001** | **0.13 (0.04)** | **<0.001** | **0.21 (0.04)** | **<0.001** | **0.19 (0.04)** | **<0.001** | **0.20 (0.04)** | **<0.001** | **0.20 (0.04)** | **<0.001** |
| Mediator×Age (older) | |  |  |  |  |  |  |  |  |  |  |  |  |  |
| AA_Horvath_ | -0.06 (0.08) | 0.496 | -0.17 (0.16) | 0.266 | **0.09 (0.04)** | **0.040** | 0.03 (0.03) | 0.455 | -0.05 (0.14) | 0.742 | -0.11 (0.16) | 0.508 | 0.09 (0.11) | 0.425 |
| AA_Hannum_ | 0.07 (0.08) | 0.392 | 0.09 (0.16) | 0.581 | **0.11 (0.04)** | **0.003** | 0.03 (0.03) | 0.382 | -0.04 (0.16) | 0.806 | -0.12 (0.20) | 0.552 | -0.10 (0.11) | 0.382 |
| AA_Pheno_ | 0.16 (0.09) | 0.056 | -0.16 (0.16) | 0.315 | **0.10 (0.04)** | **0.013** | **0.08 (0.03)** | **0.011** | 0.09 (0.18) | 0.607 | 0.03 (0.23) | 0.882 | -0.03 (0.12) | 0.827 |
| AA_Grim_ | **0.26 (0.08)** | **0.001** | -0.02 (0.16) | 0.923 | **0.10 (0.05)** | **0.035** | 0.02 (0.03) | 0.533 | 0.01 (0.16) | 0.959 | -0.19 (0.19) | 0.319 | -0.21 (0.11) | 0.055 |
| **Mediator regressed on** | |  |  |  |  |  |  |  |  |  |  |  |  |  |
| Sex (male) | 0.00 (0.03) | 0.969 | **0.08 (0.03)** | **0.016** | 0.09 (0.05) | 0.083 | **0.28 (0.03)** | **<0.001** | **0.12 (0.04)** | **0.004** | **-0.12 (0.04)** | **0.002** | -0.02 (0.03) | 0.505 |
| Age (older) | **-0.71 (0.02)** | **<0.001** | **0.39 (0.04)** | **<0.001** | **-0.33 (0.05)** | **<0.001** | **-0.11 (0.02)** | **<0.001** | **0.16 (0.05)** | **0.001** | -0.07 (0.05) | 0.177 | **-0.28 (0.06)** | **<0.001** |
| Sex×Age (older) | **0.14 (0.03)** | **<0.001** | -0.02 (0.03) | 0.488 | 0.12 (0.06) | 0.052 | -0.02 (0.04) | 0.652 | **-0.10 (0.04)** | **0.017** | -0.01 (0.04) | 0.797 | 0.07 (0.05) | 0.107 |
| B, standardised (STDYX) regression coefficient; SE, standard error; AA, epigenetic age acceleration.  ^a^ The model was controlled for zygosity.  ^b^ SEs were corrected for nested sampling.  ^c^ Significant regression coefficients at the level 0.05 are presented in bold | | | | | | | | | | | | | | |

**Mediation models in the opposite-sex twin pairs**

In the opposite-sex twin pairs, higher BMI was associated with accelerated AA_Horvath_ (B=0.24) (appendix **Table 4)**. Smoking was associated with accelerated AA_Grim_ (B=0.36) and higher sport index with slower AA_Horvath_ (B=-0.14). In all the mediation models, direct association of sex with AA_Hannum_ and AA_Grim_ was observed (B=0.22 to 0.29). When the lifestyle factors (BMI, smoking, alcohol use, and sport index) were controlled for each other in the multiple mediator models, higher BMI was still associated with accelerated AA_Horvath_ (B=0.24) and smoking with accelerated AA_Grim_ (B=0.37) (Figure 3). Surprisingly, higher sport index was associated with accelerated AA_Pheno_ (B=0.16). A positive direct effect of male sex on higher AA_Hannum_ and AA_Grim_ was observed (B=0.29 and B=0.21, respectively).

| **Appendix Table 4. Multilevel mediation models for opposite-sex twin pairs (151 pairs). Within-twin pair level standardised regression coefficients of the single mediator models are presented** | | | | | | | | | | | | | | |
| --- | --- | --- | --- | --- | --- | --- | --- | --- | --- | --- | --- | --- | --- | --- |
|  | **Mediator** ^a, b^ | | | | | | | | | | | | | |
|  | **Education** | | **Body mass index** | | **Smoking** | | **Alcohol use** | | **Sport index** | | **Leisure index** | | **Work index** | |
|  | **B (SE)** | ***p*** | **B (SE)** | ***p*** | **B (SE)** | ***p*** | **B (SE)** | ***p*** | **B (SE)** | ***p*** | **B (SE)** | ***p*** | **B (SE)** | ***p*** |
| **Epigenetic age acceleration regressed on** | | | |  |  |  |  |  |  |  |  |  |  |  |
| **Mediator** |  |  |  |  |  |  |  |  |  |  |  |  |  |  |
| AA_Horvath_ | 0.11 (0.09) | 0.213 | **0.24 (0.08)** | **0.002** | -0.04 (0.12) | 0.761 | -0.10 (0.08) | 0.209 | **-0.14 (0.06)** | **0.024** | 0.00 (0.08) | 0.957 | -0.13 (0.07) | 0.086 |
| AA_Hannum_ | 0.06 (0.08) | 0.472 | -0.05 (0.11) | 0.614 | -0.05 (0.09) | 0.571 | -0.12 (0.09) | 0.179 | 0.14 (0.08) | 0.072 | 0.11 (0.07) | 0.117 | 0.01 (0.07) | 0.895 |
| AA_Pheno_ | 0.10 (0.07) | 0.162 | 0.07 (0.09) | 0.437 | 0.11 (0.11) | 0.304 | -0.07 (0.08) | 0.376 | 0.15 (0.09) | 0.110 | 0.11 (0.07) | 0.099 | 0.04 (0.07) | 0.598 |
| AA_Grim_ | -0.15 (0.07) | 0.051 | -0.03 (0.11) | 0.808 | **0.36 (0.09)** | **<0.001** | 0.03 (0.08) | 0.687 | -0.02 (0.09) | 0.819 | -0.09 (0.07) | 0.167 | 0.08 (0.06) | 0.179 |
| **Sex (male)** |  |  |  |  |  |  |  |  |  |  |  |  |  |  |
| AA_Horvath_ | **0.12 (0.06)** | **0.049** | 0.05 (0.06) | 0.385 | 0.10 (0.06) | 0.074 | **0.13 (0.06)** | **0.033** | 0.10 (0.06) | 0.080 | 0.10 (0.06) | 0.095 | 0.11 (0.06) | 0.056 |
| AA_Hannum_ | **0.27 (0.05)** | **<0.001** | **0.27 (0.06)** | **<0.001** | **0.26 (0.06)** | **<0.001** | **0.29 (0.06)** | **<0.001** | **0.26 (0.05)** | **<0.001** | **0.28 (0.05)** | **<0.001** | **0.25 (0.05)** | **<0.001** |
| AA_Pheno_ | -0.03 (0.06) | 0.662 | -0.06 (0.06) | 0.344 | -0.05 (0.06) | 0.375 | -0.03 (0.06) | 0.687 | -0.04 (0.06) | 0.449 | -0.02 (0.06) | 0.754 | -0.09 (0.12) | 0.425 |
| AA_Grim_ | **0.22 (0.05)** | **<0.001** | **0.26 (0.06)** | **<0.001** | **0.22 (0.06)** | **<0.001** | **0.24 (0.06)** | **<0.001** | **0.25 (0.05)** | **<0.001** | **0.23 (0.05)** | **<0.001** | **0.24 (0.05)** | **<0.001** |
| **Mediator regressed on** | |  |  |  |  |  |  |  |  |  |  |  |  |  |
| Sex (male) | **-0.17 (0.06)** | **0.002** | **0.22 (0.06)** | **<0.001** | 0.07 (0.07) | 0.286 | **0.27 (0.04)** | **<0.001** | -0.01 (0.07) | 0.847 | **-0.24 (0.07)** | **0.001** | 0.07 (0.07) | 0.332 |
| B, standardised regression coefficient; SE, standard error; AA, epigenetic age acceleration. | | | | | | | | | | | | | | |
| ^a^ The model was controlled for age as twin pairs participated in measurements at slightly different ages (±1 year). | | | | | | | | | | | | | | |
| ^b^ Significant regression coefficients at the level 0.05 are presented in bold | | | | | | | | | | | | | | |

10	Muthén LK, Muthén BO. Mplus User’s Guide. Eighth Ed. Los Angeles, CA https://www.statmodel.com.
